# How can digital citizen science approaches improve ethical smartphone use surveillance among youth: traditional surveys versus ecological momentary assessments

**DOI:** 10.1101/2024.01.14.24301303

**Authors:** Sarah Al-Akshar, Sheriff Tolulope Ibrahim, Tarun Reddy Katapally

## Abstract

**Background:** Ubiquitous use of smartphones among youth poses significant challenges related to non-communicable diseases, including poor mental health. Although traditional survey measures can be used to assess smartphone use among youth, they are subject to recall bias. This study aims to compare self-reported smartphone use via retrospective modified traditional recall survey and prospective Ecological Momentary Assessments (EMAs) among youth.

**Methods:** This study uses data from the Smart Platform, which engages with youth as citizen scientists. Youth (N=436) aged 13-21 years in two urban jurisdictions in Canada (Regina and Saskatoon) engaged with our research team using a custom-built application via their own smartphones to report on a range of behaviours and outcomes on eight consecutive days. Youth reported smartphone use utilizing a traditional validated measure, which was modified to capture retrospective smartphone use on both weekdays and weekend days. In addition, daily EMAs were also time-triggered over a period of eight days to capture prospective smartphone use. Demographic, behavioural, and contextual factors were also collected. Data analyses included t-test and linear regression using SPSS statistical software.

**Results:** There was a significant difference between weekdays, weekends and overall smartphone use reported retrospectively and prospectively (p-value= <0.001), with youth reporting less smartphone use via EMAs. Overall retrospective smartphone use was significantly associated with not having a part-time job (β=0.342, 95%[CI]=0.146-1.038, p-value =0.010) and participating in a school sports team (β=0.269, 95%[CI]= 0.075-0.814, p-value=0.019). However, prospective smartphone use reported via EMAs was not associated with any behavioural and contextual factors.

**Conclusion:** The findings of this study have implications for appropriately understanding and monitoring smartphone use in the digital age among youth. EMAs can potentially minimize recall bias of smartphone use among youth, and other behaviours. More importantly, digital citizen science approaches that engage large populations of youth using their own smartphones can transform how we ethically monitor and mitigate the impact of excessive smartphone use.

**Author Summary:** Use of ubiquitous digital devices, particularly smartphones, has experienced an exponential increase among youth, a phenomenon that continues to influence youth health. Although retrospective measures have been used to understand smartphone use among youth, they are prone to measurement and compliance biases. There has been a growing interest in using ecological momentary assessments (EMAs) to assess smartphone to minimize biases associated with retrospective measures. This study uses the smart framework, which integrates citizen science, community based participatory research and systems science to ethically engage with youth citizen scientists using their own smartphones to understand smartphone use behaviours – reported by the same cohort of youth using both retrospective and prospective measures. The findings show a significant difference between smartphone use reported through retrospective and prospective EMAs, with youth reporting more smartphone use via retrospective measures. Furthermore, there were differences in contextual and behavioural factors that were associated with smartphone use reported via retrospective and prospective measures. The findings have implications for appropriately understanding and monitoring smartphone use in the digital age among youth. More importantly, digital citizen science approaches that engage large populations of youth using their own smartphones can transform how we ethically monitor and mitigate the impact of excessive smartphone use.

## Background

The prevalence of smartphone usage among the younger population has experienced a significant surge in recent years on a global scale, as evidenced by the fact that 84% of smartphone users fall within the young adult demographic. [1]. Nonetheless, there is still ambiguity regarding how youth use their smartphones and the potential implications of using ubiquitous digital tools such as smartphones [2].

In understanding smartphone use among youth, thus far, traditional self-report measures have been predominantly used [3], which pose issues such as recall bias and measurement errors [4,5]. Moreover, smartphone use includes a range of activities that encompass communication, e-learning, entertainment, and social media, among others, which are not captured by traditional survey measures [5].

Advanced population health measurement techniques such as ecological momentary assessments (EMAs), can potentially address measurement bias and capture the entire range of smartphone activities [6,7]. EMAs “capture brief, repeated assessments in natural environments with a high degree of ecological validity relative to laboratory-based investigations” [8]. EMAs are prospective measures that provide an advantage in assessing behaviours with respect to the context and the environment of the participants [9]. While traditional surveys are important in understanding behaviour retrospectively, EMAs have the advantage of minimizing recall bias and improving the validity of results [10]. Moreover, EMAs reduce the burden of responding to surveys and improve rates of compliance and feasibility for participants [11], as well as minimize social desirability bias [12].

Although current research examines the screentime of youth, including watching TV and playing video games, there is a scarcity of evidence focusing on smartphone use [13]. It is critical to appropriately understand smartphone use among youth to not only inform potential smartphone use recommendations [14], but also to determine appropriate use for digital health interventions [15]. In addition, currently no studies exist that compare retrospectively reported smartphone use using traditional survey measures and prospectively reported smartphone use using EMAs within the same cohort of youth. The objective of this study is to compare smartphone use reported via retrospective surveys and prospective EMAs by engaging with youth citizen scientists via their own smartphones. The study also investigates the demographic, behavioural, and contextual factors that are associated with smartphone use reported via retrospective surveys compared to prospective EMAs.

## Methodology

### Study Design

This study is a part of the Smart Platform, which is a citizen science and digital epidemiological initiative for ethical population health surveillance, knowledge translation, and real-time interventions [16]. Smart Platform integrates community-based participatory research and digital citizen science to ethically obtain big data from citizen-owned smartphones[16]. The study uses a combination of cross sectional and temporal longitudinal design [17]. The Research Ethics Boards at the Universities of Regina and Saskatchewan approved the Smart Platform’s application for research ethics approval (REB # 2017-029).

The research team engaged with youth citizen scientists via their smartphones over the course of eight days through the Smart Platform smartphone custom-built application. Youth citizen scientists could download the application through the iOS or Android operating systems. [17]. To understand the retrospective smartphone use behaviours, a modified traditional retrospective recall survey was deployed using a one-time triggered notification on day one, which also collected demographic characteristics, including gender, age, and socioeconomic status **(Figure 1).** The retrospective survey investigated the habits of youth citizen scientists’ smartphone use. Later, during a period of eight days, the app triggered daily EMAs to capture prospective smartphone use including weekdays and weekends **(Figure 1).** In addition, youth citizen scientists reported on their school environment, including available facilities and resources at their school, as well as the level of family and friends’ involvement and encouragement to perform physical activities[18].

**Figure 1:**
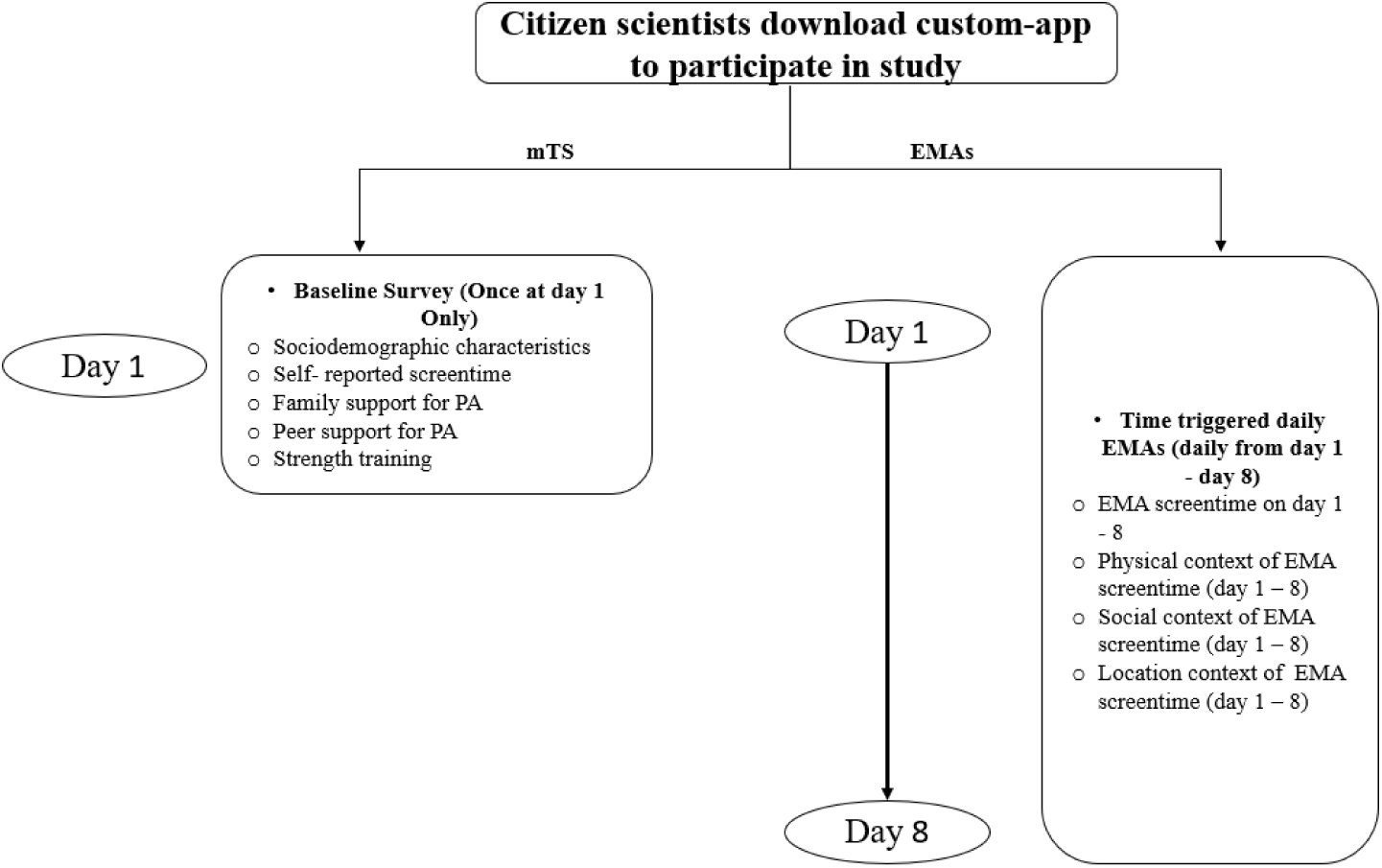
Deployment structure of prosepective and retrospective surveys

### Participants

To achieve 90% confidence level and a 5% margin of error, a sample size calculation resulted in the sample size to be at least 161. Regina Public and Catholic School Boards were contacted for recruitment of youth citizen scientists, and in-person recruitment sessions were initiated August 31^st^, 2018, until December 31^st^ of the same year. As a result, a total of 808 youth citizen scientists were recruited from five schools (aged 13-21). During the recruitment sessions, researchers demonstrated how to use the smartphone application, youth citizen scientists were able to download and join the study anytime during the recruitment period (August 31^st^ -December 31^st^, 2018). Informed consent was provided to all youth citizen scientists through the application. However, youth citizen scientists between the ages of 13-16 years were required to provide implied inform consent obtained from either their caregiver(s) or parent(s) before the scheduled recruitment sessions [18].

### Measures

#### Smartphone Use (dependent variables)

Dependent variables were mean on weekdays, weekends, and overall retrospective and prospective (EMAs) smartphone use per day. The retrospective survey questions were deployed on day one of engagement via a custom-built application, while EMAs were deployed daily (8:00 pm-11:30 pm) and were set to expire by 12:00 am.

The 9-questions Sedentary Behaviour Questionnaire Survey questions, which is a retrospective traditional data collection tool, was modified to enable the questions to be digitally deployed on citizen scientists smartphones [17]. The retrospective modified survey asked questions such as: “On a typical weekday during the school year, how much time do you spend on your smartphone for internet surfing (e.g., Facebook, Snapchat, Instagram, YouTube, Reddit, reading news, etc.)?”. From youth responses, weekday mean minutes of retrospective smartphone use was derived. Further the retrospective survey asked: “On a typical weekend during the school year, how much time do you spend on your smartphone for internet surfing (e.g., Facebook, Snapchat, Instagram, YouTube, Reddit, reading news, etc.)?”. From youth responses, weekend mean minutes of retrospective smartphone use was derived. The sum of mean weekday and weekend smartphone use was derived and averaged to obtain overall retrospective smartphone use.

To ascertain prospective smartphone use, EMA questions asked: “Which of the following did you do yesterday?” With response options to include: “Watched television”, “Internet surfing using desktop/laptop/gaming device/television screen”, “Used smartphone for internet surfing”, “Texted using a phone”, “Played games on smartphones”, “Listened to musing while sitting”, “Played a musical instrument while sitting”, “Did arts and crafts while sitting”, and “Drove or sat in a car/bus.”, “How many minutes did you spend doing this activity?”, and “With whom did you do this activity?” From these responses, weekend mean minutes of prospective smartphone use was derived. To derive overall smartphone use prospectively, a sum of weekday mean minutes of prospective smartphone use and weekend mean minutes of prospective smartphone use was derived and averaged. Both retrospective and prospective overall, weekday, and weekend mean minutes of smartphone use were the primary dependent variables of this study.

#### School PA environment (independent variable)

To capture School PA environment, the following question was asked: “Do you participate in competitive school sports teams that compete against other schools (e.g., junior varsity or varsity sports)?”, with the three response options being: “Yes”, “No” or “None offered”. Responses were dichotomized into “Disagree”, corresponding to the response options “No” or “None offered”, and “Agree”, corresponding to the response option “Yes”.

#### Physical Activity Strength training (independent variable)

The retrospective survey captured strength training using the question: “On how many days in the last 7 days did you do exercise to strengthen or tone your muscles (e.g., push-ups, sit-ups, or weight-training)?” with the eight response options being: “0 days,” “1”, “2”, “3”, “4”, “5”, “6”, or “7 days”. The responses were recoded to “less than four days”, and “four days or more”. Another question deployed to capture physical activity was “how many minutes did you spend performing moderate to vigorous physical activity on each day of the week?”.

#### Social support for PA (independent variables)

Social support for physical activity included family and friends’ support. To capture family involvement with physical activities, the following questions were asked. “How much do your parents, stepparents, or guardians encourage you to be physically active?” with the five response options being: “Strongly encourage”, “Encourage”, “Do not encourage nor discourage”, “Discourage”, or “Strongly discourage”. Responses were dichotomized into “Discourage/neutral”, which combined the response options “Do not encourage nor discourage”, “Discourage”, or “Strongly discourage”, and “Encourage”, which included the response options “Strongly encourage”, or “Encourage”. Finally, youth were asked: “How much do your parents, stepparents, or guardians support you in being physically active (e.g., driving you to team games, buying you sporting equipment)?” with the four response options being: “Very supportive”, “Supportive”, “Unsupportive”, or “Very unsupportive”. Responses were dichotomized as “Unsupportive/Neutral”, which combined the response options “Unsupportive” and “Very unsupportive”, and “supportive”, which included the response options “Very supportive” and “Supportive”.

Peer support for PA was captured by asking youth to think about their friends during the past 12 months when providing answer to this question: “How many of your closest friends are physically active?” with the six response options being: “None of my friends”, “1”, “2”, “3”, “4”, or “5 of my friends.” Responses were dichotomized as “Three or less of my friends”, corresponding to the response option “None of my friends”, “1”, “2”, “3 of my friends” and “Four or more of my friends””, corresponding to “4”, or “5 of my friends.”

#### Socio-demographic Covariates

The survey asked demographic questions that included parent education level, identity, gender, and age. Question deployed to collect information about parent education level was: “what is the highest education level for your parents?”. Responses were: “elementary”, “high school diploma”, “college”, “university”. Responses were recoded into the following categories: “below Secondary education level” corresponding to “elementary” and “Secondary and above education level” corresponding to “high school diploma”, “college”, “university”. Youth identity was captured by asking the question: “How would you describe your identity? Select all that apply” for which there were 13 response options: “First Nations”, “Dene”, “Cree”, “Metis”, “Inuit”, “African”, “Asian”, “Canadian”, “Caribbean/West Indian”, “Eastern European”, “European”, “South Asian” or “Other (please specify)”. Response options were recoded to result in the following categories due to low count values: “Indigenous Canadians” corresponding to “First Nations”, “Dene”, “Cree”, “Metis”, “Inuit”, “Non-Indigenous Canadians” corresponding to “Canadian” and “Other” corresponding to “Caribbean/West Indian”, “Eastern European”, “European”, “South Asian” or “Other. Further gender was captured by asking: “What is your gender?” which had five response options: “Male”, “Female”, “Transgender”, “Other (please specify)”, or “Prefer not to disclose”. Gender was recoded into three categories: “Female”, “Male”, or “Transgender/other (please specify)/prefer not to disclose”. Age was captured by asking citizen scientists the question “How old are you? (Age in years)”. Part-time job was captured by asking the question “do you have a part-time job?” with response options “yes” and “no”.

### Data and risk management

Data were encrypted before being saved on smartphones and sent to servers when a device established a Wi-Fi connection to protect confidentiality. The app’s permissions were controlled to avoid accessing any personal data on their smartphones (e.g., contact list or network sites visited). Media Access Control (MAC), which is based on a basic encryption method that is impossible to reverse, was used for anonymization, and to secure the information of citizen scientists. During the process of obtaining informed consent, risks and privacy management alternatives were clearly communicated to the citizen scientists [18].

### Data Analysis

Characteristics of the youth citizen scientists were reported using descriptive statistics. Pearson correlation was used to look at the association between smartphone use reported retrospectively and prospectively on weekdays and weekends. A paired sample t-test was conducted to explore if there exists a difference in mean minutes/day of retrospective smartphone use, and mean minutes/day of prospective smartphone use reported on weekday and weekend. Further, this study investigates if there exists a difference in mean minutes of smartphone use per day retrospectively and prospectively based on gender of youth citizen scientists. Multiple linear regression models were conducted to explore the contextual and behavioural factors associated with retrospective and prospective smartphone use among youth citizen scientists. Statistical analyses were conducted using IBM SPSS 29.0.0.0 Statistics Software with a significance level of p < 0.05.

## Results

A total of 808 youth citizen scientists participated in the study. However, after excluding youth citizen scientists who did not provide complete information on key dependent and independent variables, a total of 436 youth scientists constituted the final sample size. Around 56% of the youth citizen scientists were female, 5% identified as Indigenous and 64.1% did not have a part-time job (**Table 1**).

**Table 1:**
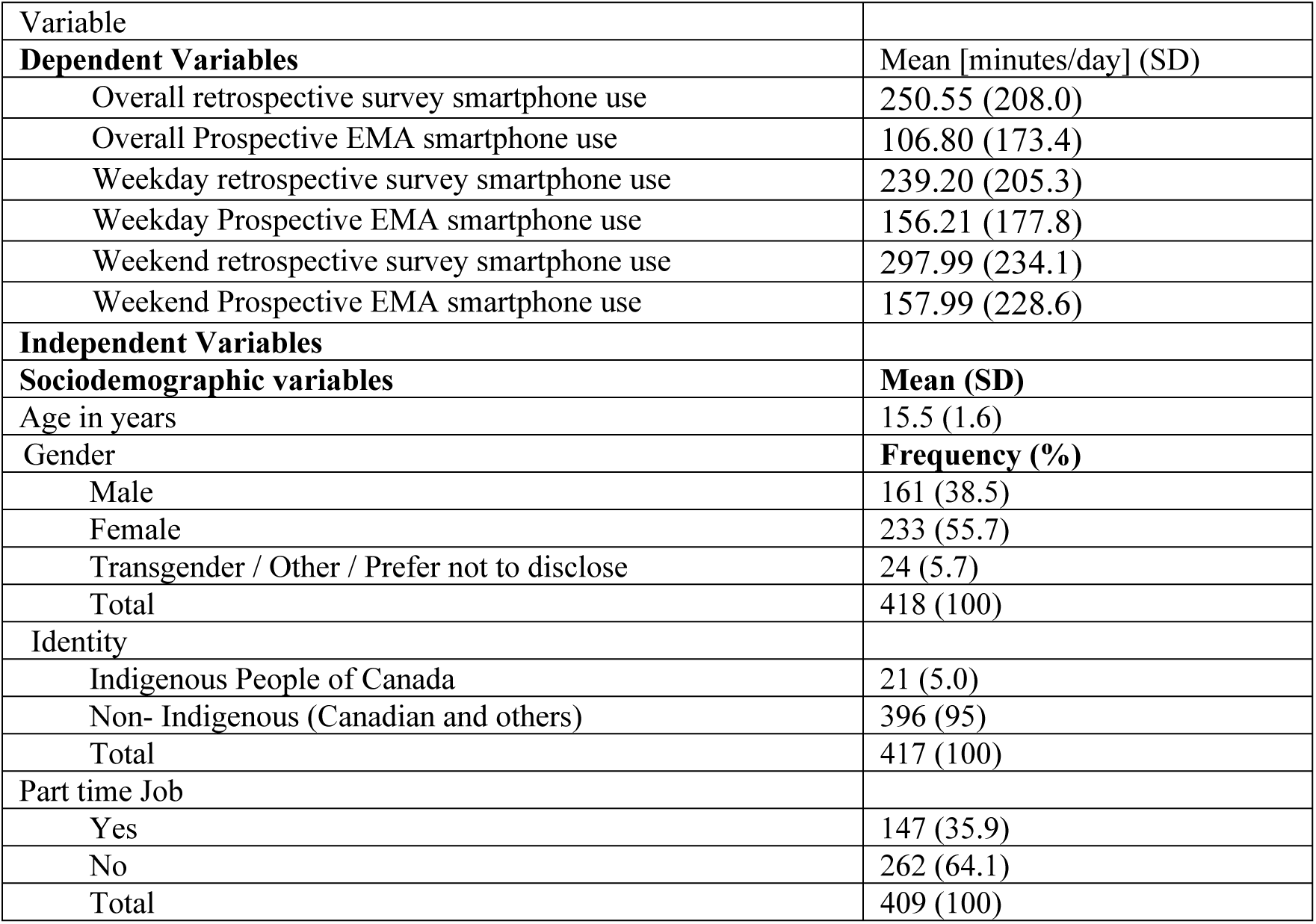
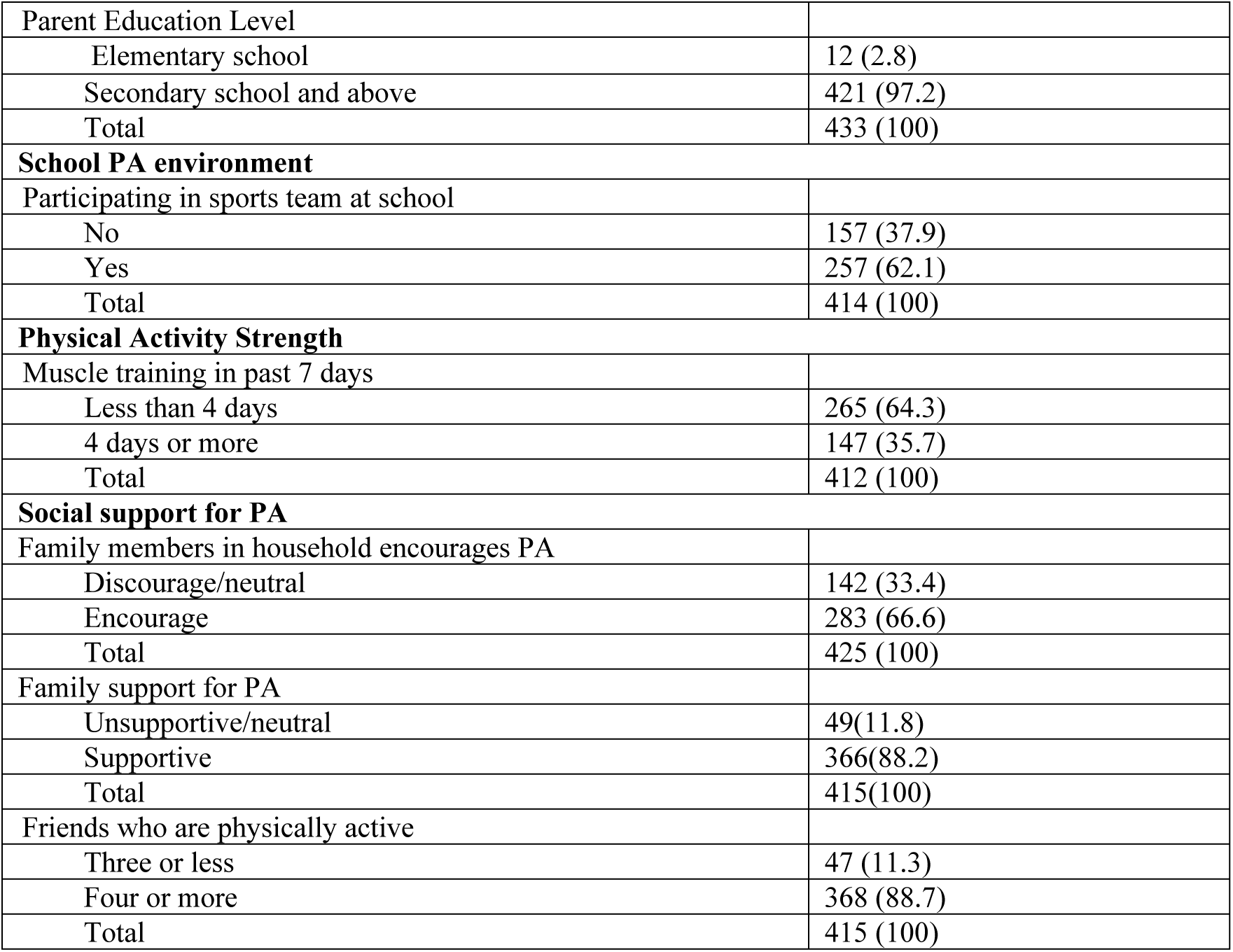
Summary of characteristics of youth citizen scientists (n=436).

There was a significant difference between overall (weekdays+weekend days) mean minutes of smartphone use/day reported retrospectively (250.55 minutes/day) and prospectively (106.80 minutes/day) (**Table 2**). Similarly, youth also reported more smartphone use on both weekdays and weekend days using retrospective surveys (239.20 minutes/day; 297.99 minutes/day), compared to smartphone use reported prospectively via EMAs (156.21 minutes/day; 157.99 minutes/day). This pattern continued among females, but not males **(Table 3).** There was a significant difference between overall (weekdays+weekend days), and weekdays mean smartphone use reported retrospectively (270 minutes, 257.92 minutes) and prospectively (88.12 minutes, 138.91) among female youth citizen scientists (p<0.001). However, there was no significant difference between weekend mean smartphone use reported retrospectively and prospectively among female citizen scientists.

**Table 2:** Paired sample t-test result showing the differences between smartphone use retrospectively and prospectively.

| Overall | Mean | t-value | df | N | p-value |
| --- | --- | --- | --- | --- | --- |
| Minutes of smartphone use reported retrospectively | 250.55 | 6.460 | 90 | 91 | <0.001 |
| Minutes of smartphone use reported prospectively | 106.80 |  |  |  |  |
| Weekday |  |  |  |  |  |
| Minutes of smartphone use reported retrospectively | 239.20 | 3.087 | 55 | 56 | <0.001 |
| Minutes of smartphone use reported prospectively | 156.21 |  |  |  |  |
| Weekend |  |  |  |  |  |
| Minutes of smartphone use reported retrospectively | 297.99 | 4.202 | 66 | 67 | <0.001 |
| Minutes of smartphone use reported prospectively | 157.99 |  |  |  |  |

**Table 3:** Paired sample t-test result showing the differences between smartphone use retrospectively and prospectively based on gender.

|  | Female |  |  |  |  | Male |  |  |  |  |
| --- | --- | --- | --- | --- | --- | --- | --- | --- | --- | --- |
| Overall | Mean | t-value | df | N | p-value | Mean | t-value | df | N | p-value |
| Overall smartphone use reported retrospectively | 270.00 | 8.308 | 58 | 59 | <0.001 | 219.44 | 1.556 | 30 | 31 | 0.130 |
| Overall smartphone use reported prospectively | 88.12 |  |  |  |  | 144.28 |  |  |  |  |
| Weekday |  |  |  |  |  |  |  |  |  |  |
| Weekday smartphone use reported retrospectively | 257.92 | 3.983 | 35 | 36 | <0.001 | 212.37 | 0.319 | 18 | 19 | 0.753 |
| Weekday smartphone use reported prospectively | 138.91 |  |  |  |  | 195.37 |  |  |  |  |
| Weekend |  |  |  |  |  |  |  |  |  |  |
| Weekend smartphone use reported retrospectively | 318.29 | 5.865 | 40 | 41 | 0.022 | 274.20 | 0.975 | 24 | 25 | 0.339 |
| Weekend smartphone use reported prospectively | 131.63 |  |  |  |  | 205.14 |  |  |  |  |

After controlling for demographic characteristics (age, gender, and identity), results of the multiple linear regression models exploring social and contextual factors associated with overall, weekday, and weekend smartphone use reported via retrospectively surveys and prospective EMAs are portrayed in tables 4, 5, and 6, respectively. Within the overall retrospective model (**Table 4 – Model 1**), youth citizen scientists who reported having a part-time job (β=0.342, 95% confidence interval [CI]=0.146, 1.038, p-value = 0.010) reported more smartphone use compared to youth citizen scientists who did not have part-time jobs. Similarly, youth citizen scientist who reported participating in a school sports team (β=0.269, 95%[CI]= 0.075, 0.814, p-value=0.019) reported more smartphone use in comparison to youth who did not participate in a school sports team. However, none of these relationships was found to be significant in the prospective EMA model (**Table 4-Model 2**).

**Table 4:** Linear Regression models of overall smartphone use reported retrospectively vs. prospectively.

| Variables | Retrospective (Model 1) |  | Prospective (Model 2) |  |
| --- | --- | --- | --- | --- |
|  | Standardized Coefficients Beta | (95% CI) p-value | Standardized Coefficient Beta | (95% CI) p-value |
| (constant) |  | (-0.856-5.221)<br>0.156 |  | (-0.402-9.789)<br>0.070 |
| **Sociodemographic variables** |  |  |  |  |
| Parent Education level: Below secondary school (Ref) |  |  |  |  |
| Secondary school and above | 0.175 | (-0.063-0.678)<br>0.103 | 0.149 | (-0.231-1.009)<br>0.214 |
| Part-time job: Yes (Ref) |  |  |  |  |
| Not having a part time job | 0.342 | **(0.146-1.038)**<br>**0.010\*\*\*** | -0.010 | (-0.780-0.731)<br>0.948 |
| **Physical Activity Strength** |  |  |  |  |
| Moderate to vigorous physical activity per day | 0.222 | (-0.017-0.475)<br>0.068 | 0.166 | (-0.155-0.667)<br>0.218 |
| Muscle training: less than 4 days (Ref) |  |  |  |  |
| Muscle Training- 4 days or more | 0.129 | (-0.145-0.568)<br>0.241 | -0.214 | (-1.119-0.076)<br>0.086 |
| **School PA environment** |  |  |  |  |
| Participating in a sports team: No (Ref) |  |  |  |  |
| Participating in a sports team | 0.269 | **(0.075-0.814)**<br>**0.019\*\*\*** | -0.076 | (-0.806-0.430)<br>0.546 |
| **Family and Friends involvement in PA** |  |  |  |  |
| Family support for physical activity: Unsupportive/neutral (Ref) |  |  |  |  |
| Family support for physical activity: Supportive | 0.221 | (-0.021-0.976)<br>0.060 | 0.035 | (-0.719-0.943)<br>0.789 |
| Family encouragement for physical activity: Discourage/neutral (Ref) |  |  |  |  |
| Family encouragement for physical activity: Encourage | -0.062 | (-0.517-0.295)<br>0.586 | -0.140 | (-1.054-0.313)<br>0.283 |
| Number of physically active friends: Three or less (Ref) |  |  |  |  |
| Four or more physically active friends | -0.046 | (-0.444-0.299)<br>0.698 | 0.050 | (-0.509-0.743)<br>0.711 |
| *** $p < 0.05$ all regression models controlled for: age, gender, and identity | | | | |

**Table 5:** Linear regression models of weekday smartphone use reported retrospectively vs. prospectively.

|  | Retrospective (Model 3) |  | Prospective (Model 4) |  |
| --- | --- | --- | --- | --- |
|  | Standardized Coefficients<br>Beta | (95% CI)<br>P-value | Standardized Coefficient<br>Beta | (95% CI)<br>P-value |
| (constant) |  | (-1.305-4.890)<br>0.252 |  | (-1.588-10.778) 0.140 |
| **Sociodemographic variables** |  |  |  |  |
| Parent Education level: Below secondary school (Ref) |  |  |  |  |
| Secondary school and above | 0.233 | **(0.030-0.785)**<br>**0.035\*\*\*** | 0.135 | (-0.398-0.983) 0.396 |
| Part-time job: Yes (Ref) |  |  |  |  |
| Not having a part time job | 0.313 | **(0.086-0.995)**<br>**0.020\*\*\*** | 0.150 | (-0.490-1.111) 0.437 |
| **Physical Activity Strength** |  |  |  |  |
| Moderate to vigorous physical activity per day | 0.158 | (-0.088-0.414)<br>0.200 | 0.218 | (-0.209-0.805) 0.241 |
| Muscle training: less than 4 days (Ref) |  |  |  |  |
| Muscle Training- 4 days or more | 0.150 | (-0.118-0.609)<br>0.182 | -0.204 | (-1.176-0.307) 0.242 |
| **School PA environment** |  |  |  |  |
| Participating in a sports team: No (Ref) |  |  |  |  |
| Participating in a sports team | 0.197 | (-0.053-0.701)<br>0.090 | -0.044 | (-0.804-0.622) 0.798 |
| **Family and friends' involvement in PA** |  |  |  |  |
| Family support for physical activity: Unsupportive/neutral (Ref) |  |  |  |  |
| Family support for physical activity: Supportive | 0.246 | **(0.021-1.037)**<br>**0.042\*\*\*** | -0.025 | (-1.082-0.933) 0.882 |
| Family encouragement for physical activity: Discourage/neutral (Ref) |  |  |  |  |
| Family encouragement for physical activity: encourage | -0.073 | (-0.543-0.285)<br>0.535 | -0.187 | (-1.306-0.431) 0.314 |
| Number of physically active friends: Three or less (Ref) |  |  |  |  |
| Four or more physically active friends | -0.088 | (-0.516-0.240)<br>0.469 | -0.039 | (-0.867-0.702) 0.832 |
| *** $p < 0.05$ all regression models controlled for: age, gender, and identity | | | | |

In the weekday retrospective regression model (**Table 5 – Model 3**), youth who reported their parents had attained education levels at Secondary school and above (β= 0.223, 95%[CI]= 0.030, 0.785, p-value=0.035) were significantly associated with more minutes of smartphone use compared to youth whose parents had only elementary school education. Similarly, youth who reported having family support for physical activity (β=0.246, 95%[CI]= 0.021, 1.037, p-value=0.042) reported more smartphone use, compared to youth who do not have family support for physical activity. In addition, youth who did not have a part-time job (β= 0.313, 95%[CI]= 0.086, 0.995, p-value=0.020) were significantly associated with more weekday smartphone use compared to youth who had a part-time job. These associations were not found to be significant in the prospective model (**Table 5 – Model 4**)

In the weekend retrospective model (**Table 6 - Model 5**), youth citizen scientists who did not have a part-time job (β=0.333, 95%[CI]= 0.133, 1.182, p-value=0.015) reported significantly greater smartphone use in comparison with had a part-time job. Similarly, youth who reported participating in a school sports team was found to be associated with more smartphone use (β=0.261, 95%[CI]= 0.057, 0.927, p-value=0.027) as compared to youth who did not report participating in school sports team. However, these significant associations were not found in the prospective EMA model (**Table 6 – Model 6**).

**Table 6:** Linear regression models of weekends smartphone use reported retrospectively vs. prospectively.

|  | **Retrospective (Model 5)** |  | **Prospective (Model 6)** |  |
| --- | --- | --- | --- | --- |
|  | Standardized Coefficients Beta | (95% CI) p-value | Standardized Co efficient Beta | (95% CI) p-value |
| (constant) |  | (-2.463-4.686) 0.537 |  | (-2.055-10.685) 0.180 |
| **Sociodemographic variables** |  |  |  |  |
| Parent Education level: Below secondary school (Ref) |  |  |  |  |
| Secondary school and above | 0.122 | (-0.192-0.680) 0.268 | 0.147 | (-0.328-1.050) 0.298 |
| Part-time job: Yes (Ref) |  |  |  |  |
| Not having a part time job | 0.333 | **(0.133-1.182)**<br>**0.015\*\*\*** | -0.115 | (-1.197-0.604)<br>0.511 |
| **Physical Activity Strength** |  |  |  |  |
| Moderate to vigorous physical activity per day | 0.220 | (-0.030-0.550) 0.078 | 0.209 | (-0.151-0.711)<br>0.197 |
| Muscle training: less than 4 days (Ref) |  |  |  |  |
| Muscle Training- 4 days or more | 0.099 | (-0.235-0.605) 0.382 | -0.156 | (-1.034-0.330)<br>0.304 |
| **School PA environment** |  |  |  |  |
| Participating in a sports team: No (Ref) |  |  |  |  |
| Participating in a sports team | 0.261 | **(0.057-0.927)**<br>**0.027\*\*\*** | -0.088 | (-0.870-0.466)<br>0.547 |
| **Family and Friends involvement in PA** |  |  |  |  |
| Family support for physical activity: Unsupportive/neutral (Ref) |  |  |  |  |
| Family support for physical activity: Supportive | 0.166 | (-0.177-0.996) 0.168 | 0.025 | (-0.852-0.989)<br>0.881 |
| Family encouragement for physical activity: Discourage/neutral (Ref) |  |  |  |  |
| Family encouragement for physical activity: encourage | -0.043 | (-0.566-0.389) 0.713 | -0.116 | (-0.973-0.424)<br>0.433 |
| Number of physically active friends: Three or less (Ref) |  |  |  |  |
| Four or more physically active friends | 0.015 | (-0.410-0.463) 0.904 | -0.061 | (-0.830-0.570)<br>0.711 |
| *** $p < 0.05$ all regression models controlled for: age, gender, and identity | | | | |

## Discussion

The objective of this study was to compare smartphone use reported via retrospective surveys and prospective EMAs by engaging with youth citizen scientists via their own smartphones. The study also investigated the behavioural, and contextual factors that are associated with smartphone use reported via retrospective surveys and prospective EMAs. The primary findings of this study show that there is a significant difference in smartphone use reported retrospectively (via validated surveys) and prospectively (via EMAs). While evidence generally indicates that retrospective data collection is prone to bias irrespective of over- or under-estimation [19–21], to our knowledge, no research has been carried out to compare retrospective recall surveys and prospective EMAs to assess smartphone use within the same cohort of participants.

While there is some evidence of over-estimation of sedentary behaviours when reporting retrospectively [19], and discrepancies between reporting smartphone use between males and females [22], our study is the first digital epidemiological investigation that compared smartphone use reporting retrospectively and prospectively among the same cohort of youth. There is some evidence that individuals who are more socially engaged might over-estimate their smartphone use, while individuals who under-estimate their smartphone use may have lost track of time [20]. In general, evidence indicates that prospective measurement of behaviours using EMAs is more accurate due to less susceptibility to recall bias [17,23].

Although the World Health Organization guidelines indicate that youth aged 15-18 should not spend more than two hours of screentime per day [24], current evidence indicates that youth across the world spend significantly more time than two hours/day on screens [25,26]. The uncontrolled screentime has detrimental effects on both physical and mental health leading to increased risk of non-communicable chronic diseases such as obesity, diabetes, heart diseases, reduced sleep quality as well as stress, anxiety, and depression [27]. More importantly, the screentime guidelines do not specify the time that youth should spend on mobile devices, particularly smartphones, which most youth have access to in this digital age [28,29]. This gap in guidelines needs to be addressed, particularly because smartphones are the primary devices of day-to-day functioning, yet ironically excessive smartphone use is also associated with poor youth health outcomes [4,14,30] However, to understand associations between smartphone use and youth health outcomes, it is imperative to obtain valid and reliable data, which, again ironically is possible via smartphones themselves due to their near universal usage among youth [4,31]. This study utilized the Smart platform [18], a digital citizen science platform that engaged youth as citizen scientists to obtain both retrospective and prospective smartphone use via their own smartphones [6].

This approach also enabled our team to obtain relevant data on behavioral, contextual, and demographic factors to investigate the association of these variables with both retrospective and prospective smartphone use among youth. Moreover, as evidence indicates that smartphone use and screentime in general varies between weekdays and weekend days [4,30] this study explored the association of behavioural and contextual factors with overall (weekday+weekend day) youth smartphone use, as well as weekday and weekend day smartphone use reported by both retrospective surveys and prospective EMAs. One consistent finding was that there were no significant associations between socio-demographic and contextual factors with overall, weekday, and weekend day smartphone use reported via EMAs.

However, retrospective models showed some common associations between sociodemographic and contextual factors with overall, weekday and weekend day smartphone use. Youth who reported that they did not have a part-time job also reported significantly more overall, and weekend day smartphone use in comparison with youth who had a part-time job - a finding that is consistent with some existing evidence [33], and potentially an indication that youth, due to the nature of part-time work they do, do not have access to smartphones consistently.

Interestingly, youth who participated in sports teams also reported greater overall and weekend day smartphone use. This finding contradicts existing evidence that shows lower smartphone use among students that participate in school sports teams [34]. However, some studies also report increased social media engagement among students during school sporting events, which may explain increased smartphone use association with participation in school sports teams [35].

The findings of this study also showed that youth whose parents had at least secondary school education reported higher smartphone use on weekdays, another contradictory finding because current evidence indicates that higher parental education is associated with lower smartphone use among children and youth [36]. Likewise, youth who reported family support for physical activity also reported higher smartphone use, another unexpected association that could potentially be attributed to social desirability bias, where individuals report more support for physical activity [37].

Irrespective of the direction of association of contextual and sociodemographic variables, there are some important aspects that need to be highlighted in understanding smartphone use in general, and in particular among youth. First, all significant findings were depicted in the retrospective smartphone use reporting models, and no significant findings were depicted in the prospective smartphone use reporting (EMAs). This in itself is a major finding that emphasizes the rationale for conducting this study – it is critical that more accurate measures of reporting be standardized to understand smartphone use because retrospective self-reporting data collection tools are prone to bias and misclassification [14,32,38].

Second, it is imperative to appreciate the complexity of smartphone use, which includes various behaviours and motivations, such as social media use, gaming, and texting, among others, which have varying associations with health outcomes [40–43]. To capture these effectively and accurately, it is critical to move towards prospective measures such as EMAs that minimize recall bias, particularly when the reporting is complicated by the range of behaviours [44]. Objective measures provide accurate overall smartphone use estimates [6], however, to capture the variation of smartphone use ethically, while preserving privacy can be challenging [45]. Finally, the contextual and sociodemographic associations of smartphone use also have to be captured prospectively to minimize bias, while again, appreciating the complexity of smartphone access itself [11]. For instance, youth from affluent households with more educated parents might have easier and earlier access to smartphones, which could in itself contradict accepted patterns that associate higher socioeconomic status with more positive health behaviours and outcomes [46,47].

Perhaps more importantly, the approaches we use to capture digital data ethically from citizens need to be revisited. In scientific research settings, power often lies with those who have knowledge such as researchers, which has been an obstacle for meaningful data collection, particularly if the data are sensitive and need to be obtained via citizen devices [48]. Digital citizen science is an approach that can democratize science through engagement with citizens ethically and directly to obtain prospective big data [49,50]– an approach that was utilized in collected data from youth in this study via their own smartphones [6,51].

Appropriate measurement of smartphone use among youth is critical to not only inform potential smartphone use recommendations [14], but also to determine appropriate use for digital health interventions [15] that have the potential to address mental health illnesses at reduced cost and improve access to resources [31], a measurement that could potentially be improved via digital citizen science approaches [14,18].

## Strengths and limitations

No studies have compared smartphone use among youth using validated retrospective traditional surveys and prospective EMAs. The strengths of this study include utilizing citizen science approaches, where youth used their own devices to engage with the researchers. The key limitation is that the smartphone use reported in this study does not delineate between the different types of smartphone use informed by varied motivations – social media, texting, gaming, among others. Another limitation is that data were collected during one season even though there is evidence that screentime and sedentary behaviours are influenced by changes in weather [56,57]. Future studies should not only measure different types of smartphone use, but also aim to capture objective data to improve accuracy of reporting, while taking seasonality into consideration [58].

## Conclusion

The findings of this study have implications for appropriately understanding and monitoring smartphone use in the digital age among youth. EMAs can potentially minimize recall bias of smartphone use among youth, and other behaviours. More importantly, digital citizen science approaches that engage large populations of youth using their own smartphones can transform how we ethically monitor and mitigate the impact of excessive smartphone use.

## Data Availability

The data that underpins the results of this study can be accessed through the following link: https://figshare.com/articles/dataset/RetrtoVsPro_Dataset_xlsx/24969579

